# External validation and updating of a prediction model for the diagnosis of gestational diabetes mellitus

**DOI:** 10.1101/2021.12.05.21267329

**Authors:** Shamil D. Cooray, Kushan De Silva, Joanne Enticott, Shrinkhala Dawadi, Jacqueline A. Boyle, Georgia Soldatos, Eldho Paul, Vincent Versace, Helena J. Teede

## Abstract

**Introduction:** The Monash early pregnancy prediction model calculates risks of developing GDM and is internationally externally validated and implemented in practice, however some gaps remain.

**Objective:** To validate and update Monash GDM model, revising ethnicity categorisation, updating to recent diagnostic criteria, to improve performance and generalisability.

**Methods:** Routine health data for singleton pregnancies from 2016 to 2018 in Australia included updated GDM diagnostic criteria. The Original Model predictors were included (age, body mass index, ethnicity, diabetes family history, past-history of GDM, past-history of poor obstetric outcomes, ethnicity), with ethnicity revised. Updating model methods were: recalibration-in-the-large (Model A); re-estimation of intercept and slope (Model B), and; coefficients revision using logistic regression (Mode1 C1 with original eight ethnicity categories, and Mode1 C2 with updated 6 ethnicity categories). Analysis included ten-fold cross-validation, performance measures (c-statistic, calibration-in-the-large value, calibration slope and expected-observed (E:O) ratio) and closed testing examining log-likelihood scores and AIC compared models.

**Results:** In 26,474 singleton pregnancies (4,756, 18% with GDM), we showed that temporal validation of the original model was reasonable (*c*-statistic 0.698) but with suboptimal calibration (E:O of 0.485). Model C2 was preferred, because of the high c-statistic (0.732), and it performed significantly better in closed testing compared to other models.

**Conclusions:** Updating of the original model sustains predictive performance in a contemporary population, including ethnicity data, recent diagnostic criteria, and universal screening context. This supports the value of risk prediction models to guide risk-stratified care to women at risk of GDM.

**Trial registration details:** This study was registered as part of the PeRSonal GDM study on the Australian and New Zealand Clinical Trials Registry (ACTRN12620000915954); Pre-results.

## Introduction

Gestational diabetes mellitus (GDM), defined as glucose intolerance that develops during pregnancy^1^ and is increasing in prevalence. Risk prediction modelling for GDM, undertaken in early pregnancy, offers clinical utility in several scenarios. These include in GDM prevention, by identifying and targeting lifestyle intervention for those at high risk, with 24% reduction in GDM prevalence reported in a meta-analysis of individual participant data.^2^ In GDM screening and diagnosis, recommended to enable treatment and reduce pregnancy risks^3,4^, risk prediction can optimise selective screening by recognising the cumulative and interdependent effects of individual risk factors. Furthermore, an early pregnancy prediction model for the diagnosis of GDM allows personalised risk assessment approaches and risk-stratified pregnancy care with targeted delivery of more intensive monitoring and treatment to those at highest risk.^5^ In this context, early pregnancy clinical prediction models have been developed to predict the diagnosis of GDM.^6^ However, these models require external validation in new settings to demonstrate transportability and are rarely updated to optimise performance for these settings.

The Original 2011 Monash Model (hereafter referred to as the Original Model) was designed to be used in early pregnancy and is now one of multiple available screening tools.^6,7^ Uniquely, it has been independently, externally validated and demonstrated to perform well across diverse populations internationally, across variable diagnostic approaches and criteria.^8 9^ Recently, the model has also been updated to include blood glucose levels and clinical utility has been demonstrated using reclassification analysis.^10^ It is now applied in clinical practice, facilitated by an accessible simple online tool, especially in the context of reducing oral glucose tolerance tests to those at highest risk to reduce COVID-19 viral transmission exposure at testing.^11^

Remaining gaps for the Original Model include external validation and updating in settings where universal screening for GDM is undertaken and diagnosis made using the prevailing International Association of Diabetes and Pregnancy Study Groups (IADPSG) diagnostic criteria.^12^. Since the original model was developed, and with altered diagnostic criteria, GDM has increased with up to a doubling in prevalence,^13^ without improved outcomes and there is a need to identify those women within this cohort at highest risk.^14^ Also, given ethnicity is a well-established risk factor for the diagnosis of GDM,^15^ there is a need to include generalisable ethnicity variables particularly in a multicultural settings such as in Australia. Therefore, here we aimed to externally validate, and if necessary, update the Original Model to predict GDM development applying IADPSG criteria and universal screening. We also aimed to explore the impact of revised ethnicity categories on model performance. Finally we aimed to explore and compare the different approaches to updating risk prediction models to advance the broader risk prediction field.

## Methods

Here we build on the Original Model that was developed, internally validated, subsequently internationally externally validated and applied clinically. Methods and reporting were guided by the Transparent Reporting of a multivariable prediction model for Individual Prognosis or Diagnosis (TRIPOD) statement.^16^ We address gaps in model generalisability by using a contemporary population diagnosed with GDM on IADPSG criteria. We performed the following steps:

1. Evaluate the performance of the Original Model in a contemporary population applying IADPSG criteria and universal screening.
2. Update the Original Model exploring a range of methods available to update logistic regression-based models. These methods incrementally increase the degree of change made to the original internationally validated model. The first two methods (A and B) are simple recalibration methods; the third method includes re-estimation of all regression coefficients of the original model:
  a. Recalibration-in-the-large creating - Model A
  b. Re-estimation of intercept and slope creating - Model B
  c. Revision of all coefficients using logistic regression:
    i. including the previous 8 ethnicity categories - Model C1
    ii. including revised ethnicity 6 categories - Model C2
3. Finally, a closed testing procedure compared the updated models A, B, C1 and C2 to investigate the method that may best balance performance improvements with extensiveness of model revision.^17^ Model performance was evaluated, in terms of discrimination (concordance statistic [*c*-statistic]) and calibration (calibration-in-the-large, calibration slope and calibration plot).

Study registered as part of the PeRSonal GDM study (ACTRN12620000915954).^18^

### Original Model for the prediction of GDM diagnosis in early pregnancy

The development and internal validation of the Original Model is previously described.^7^ Briefly, a dataset of 2880 singleton pregnancies who delivered at Monash Medical Centre, Melbourne, Australia in 2007 was used to develop the model and internally validated. Dichotomous outcome was a diagnosis of GDM using modified WHO criteria following the prevailing recommendations of the Australasian Diabetes in Pregnancy Society.^19^ The final model included six predictors: age, body mass index (BMI), ethnicity, family history of diabetes, past history of GDM, and poor obstetric outcome. Age and BMI were categorical variables using five age categories and six BMI categories respectively. Ethnicity was classified using an eight-category classification system based on country of birth. The model was presented as a simplified point scoring system, and since was independently externally validated demonstrating sustained performance across diverse populations internationally.^8-10^

### The 2021 validation and updating dataset

We used routinely collected health data for singleton pregnancies resulting in a birth from 1 January 2016 to 31 December 2018 at Monash Health. Monash Health is Australia’s largest health service and includes three maternity hospitals serving an ethnically diverse population in Melbourne within a universal and freely accessible health system. Maternity, birth, and neonatal data are collected and reported during routine care. We deterministically linked pathology data and clinical data extracted from the medical record. The data source details are described elsewhere.^20^ Characteristics of the original 2011 development dataset are compared to the 2021 validation and updating dataset in **Error!** Reference source not found..

**Figure 1:**
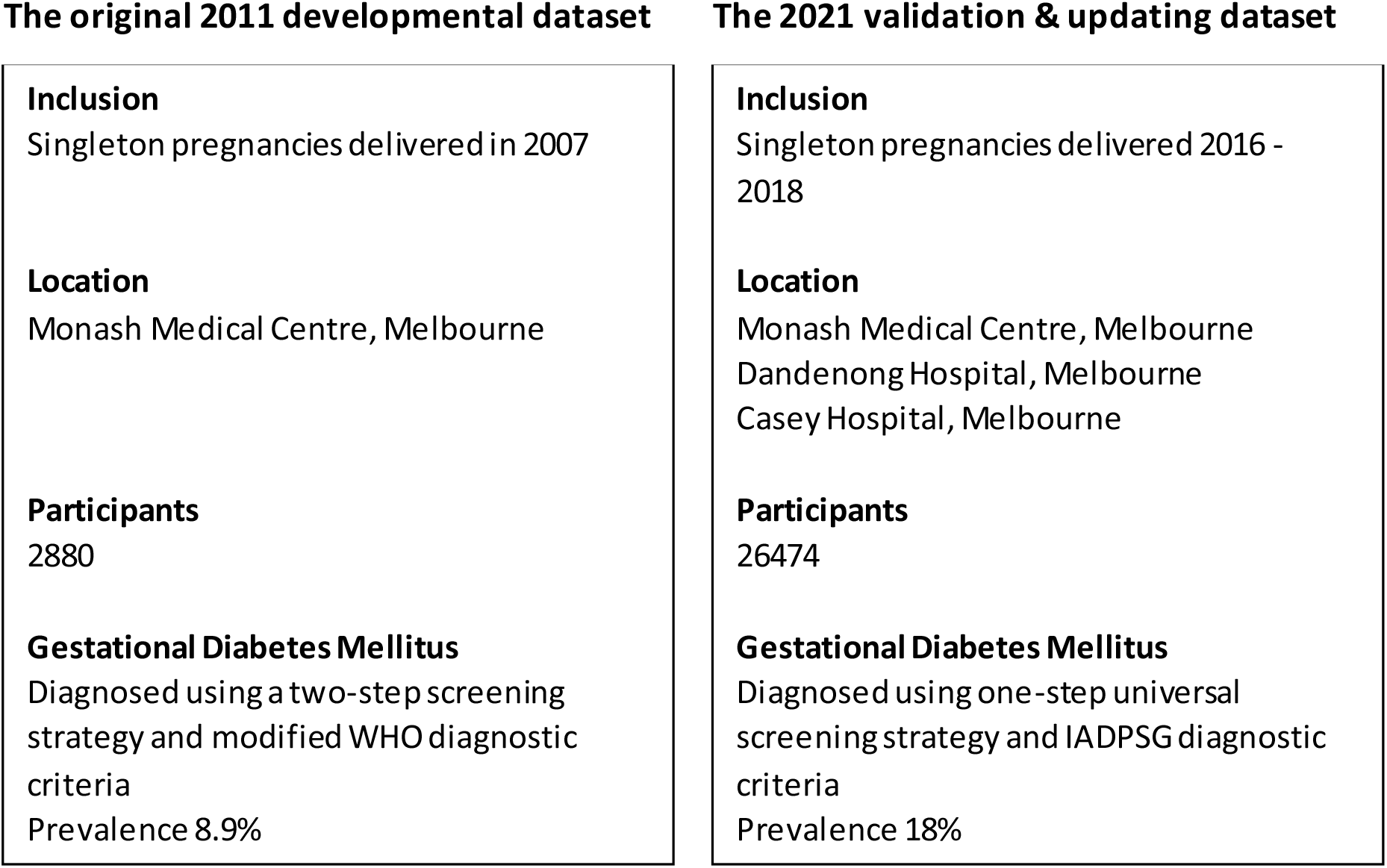
Schematic presentation of the characteristics of the original 2011 development dataset and the 2021 validation dataset. Abbreviations: IADPSG, International Association of Diabetes and Pregnancy Study Groups. Schema format adapted from Janssen et al., (2008).^21^

### Missing data and imputation

The Original Model was developed using complete case analysis. However, this approach can lead to bias and a loss of power.^22^ Therefore, we employed contemporary methods and imputed missing values. Missingness patterns of each variable was assessed and deemed to be consistent with missing at random or missing completely at random assumptions. Thereafter, missing data were multiply imputed using chained equations; ordered logistic regressions for ordinal categorical variables, multinomial logistic regression for nominal multi-level (>2 levels) categorical variables, and binary logistic regressions for dichotomous categorical variables. Sensitivity analysis of imputation was performed by comparing distributions of variables in original and complete datasets.

### Outcome definition

Diagnosis of GDM was defined using the International Association of Diabetes and Pregnancy Study Groups (IADPSG) diagnostic criteria in a one-step universal screening strategy.^12,23^

### Statistical analysis

#### Step 1: Evaluating performance of the Original Model via temporal validation

We evaluated the performance of the Original Model by applying it to a contemporary validation dataset (temporal validation) with GDM. If the Original Model did not perform as well in the contemporary validation as originally, then this would support model updating.^24^

#### Step 2: Methods to recalibrate the Updated Models

We undertook recalibration of the Original Model following three independent model updating methods.^25^

#### Step 2a: Model A (Recalibration-in-the-large)

Recalibration-in-the-large is a calibration method that is useful when outcome frequency has changed as in this case. The baseline risk of the Original Model is adjusted (i.e. adjusted intercept) to the new dataset. It reflects the differences between the setting in which the model was developed and that to which it is being applied.^24^ We estimate a single recalibration factor to correct the average of all predictions effectively shifting the intercept of the calibration plot while the original calibration slope remains unadjusted.^26^ As only a single parameter of the Original Model is updated, it is a logical first step.

#### Step 2b: Model B (Re-estimation of intercept and slope)

An updating model method where the intercept and calibration slope from the Original Model are re-estimated. All predictor effects are updated with a single overall correction factor which may potentially account for the new diagnostic criteria, using the method described by Janssen and colleagues.^26^

#### Step 2c: Models C1 and C2 (Revision of all coefficients using logistic regression)

Complete recalibration by revising regression coefficients for all predictor variables using fixed effects multiple logistic regression. We did this twice using (1) the original ethnicity 8 classifications, and (2) new ethnicity 6 reclassifications (Supplementary Table S1). The ethnicity classification system was updated to reflect international ethnicity categories with ethnicity self-reported. This differs to the Original Model where ethnicity was extrapolated from country of birth and was intended to improve accuracy and enhance generalisability to other settings. The updated classification system was aligned to the Australian Standard Classification of Cultural and Ethnic Groups.^27^ Missing ethnicity were deduced from country of birth and preferred language. This method enables the ethnicity classification system to be updated with a system that is less overfitted to the development dataset and more generalisable to new populations. With this large new dataset from an ethnically diverse population, we had a low likelihood of overfitting. This differs from *de novo* model development because the predictor selection is based on the original model development study.

#### Step 3: Comparing models

Predictive performance measures and a closed testing procedure were examined to select the best model. The performance of the Original Model was temporally validated. Updated Models A, B, C1 and C2 were evaluated using ten-fold cross-validation.

Discrimination, the ability of a model to distinguish a patient with the outcome from those without.^28^ was reported using the *c*-statistic (value 0–1, discriminative if > 0.5).^28^

Calibration examines agreement between predicted and observed risks of GDM. We report calibration slopes, calibration-in-the-large values and calibration plots. A calibration slope summarises agreement between predicted and observed risks (value 0–1: values near 1 show accuracy and values < 1 suggest predicted risks that are too extreme).^28^ Calibration-in-the-large indicates the extent that predictions are systematically too low or too high, with predicted risks under-estimated if > 0 or over-estimated if < 0).^28^ A calibration plot compares predictions (x-axis) with observed risk (y-axis) with perfect predictions lying on 45° reference line.

As noted by others^17^, particularly in small samples, model performance estimates are often too optimistic when model coefficients are revised using data that wasn’t applied in the original model development. Demonstrated in samples with n=278, n=822, and n=2,019 (and larger simulated samples up to n=2,180) the utility of a closed testing procedure to identify the best model less affected by overfitting.^17^ It proposes to investigate the update method that may balance the evidence for more extensive updating methods in the validation dataset with the danger of overfitting particularly when small samples sizes are available.^17^ As our sample was large (>25,000), collected from a diverse multicultural population, we supplemented the routine measures of model performance with this closed testing procedure. To do this, each of the four updated models were systematically compared on model fit with a pre-specified statistical significance level alpha (set at 0.001 to account for multiple comparisons).

Analyses used Stata version 16.1 (College Station, Texas, USA: StataCorp LLC) and R version 3.6.2 (Vienna, Austria: R Core Team).

## Results

The validation dataset consisted of 26,474 singleton pregnancies.

### Characteristics of participants

The sample characteristics are in Table 2**Error! Reference source not found**.. Prevalence of GDM in the new dataset was 18.0% (4,756/26,474). Mean age and BMI of participants in the imputed sample of the validation dataset were 30.4 ± 5.2 years and 26.4 ± 6.2 kg/m^2^, and 42.1% of women had a family history of diabetes, 6.9% past-history of GDM, and 9.0% history of poor obstetric outcome. Notably, a diagnosis of GDM, past-history of GDM, and a history of poor obstetric outcome was more common in the new dataset compared to the Original Model development dataset.

**Table 1.**
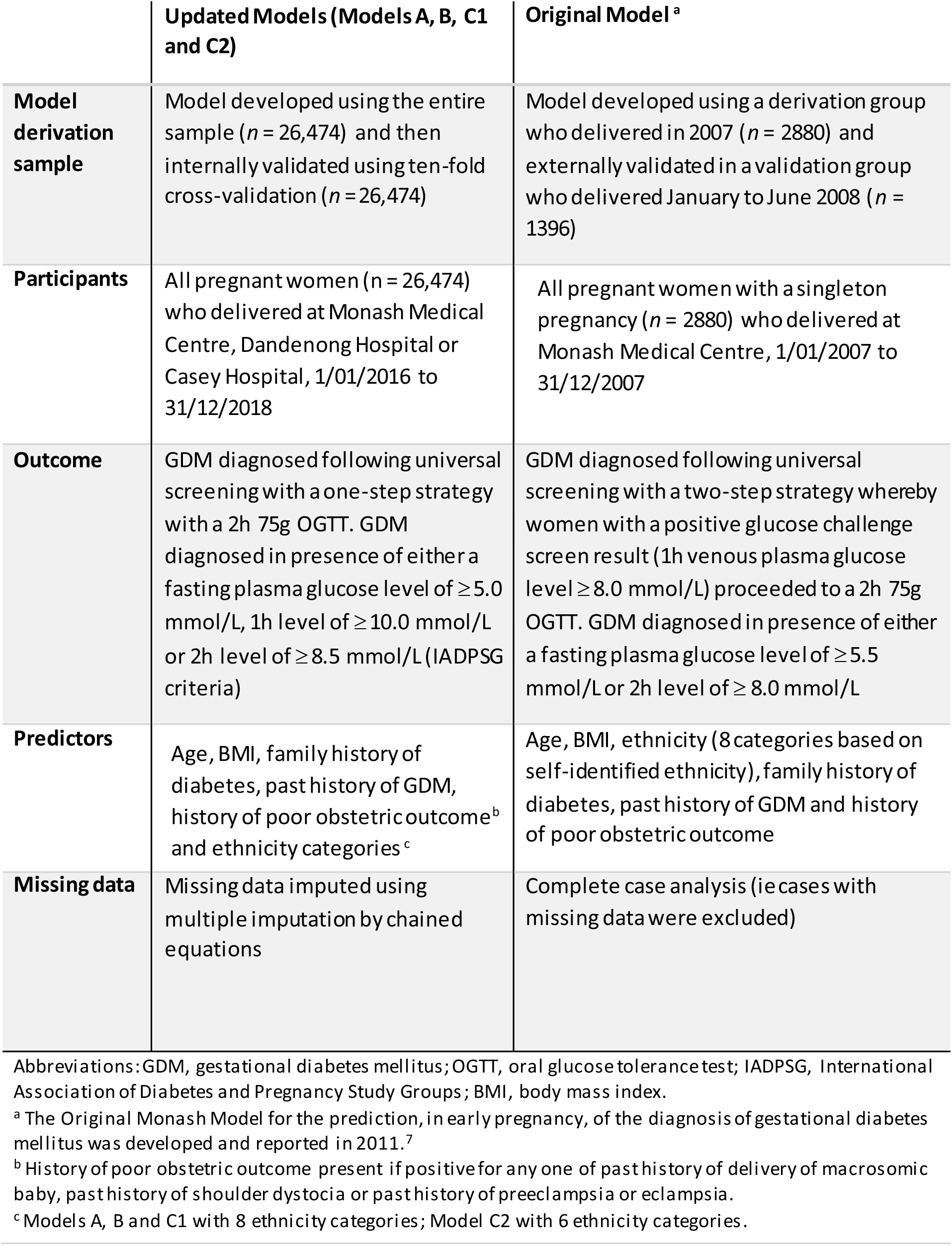
The Updated Models compared to the Original Model in terms of population, outcome, predictors and statistical analysis methods.

|  | Updated Models (Models A, B, C1 and C2) | Original Model <sup>a</sup> |
| --- | --- | --- |
| **Model derivation sample** | Model developed using the entire sample ($n = 26,474$) and then internally validated using ten-fold cross-validation ($n = 26,474$) | Model developed using a derivation group who delivered in 2007 ($n = 2880$) and externally validated in a validation group who delivered January to June 2008 ($n = 1396$) |
| **Participants** | All pregnant women ($n = 26,474$) who delivered at Monash Medical Centre, Dandenong Hospital or Casey Hospital, 1/01/2016 to 31/12/2018 | All pregnant women with a singleton pregnancy ($n = 2880$) who delivered at Monash Medical Centre, 1/01/2007 to 31/12/2007 |
| **Outcome** | GDM diagnosed following universal screening with a one-step strategy with a 2h 75g OGTT. GDM diagnosed in presence of either a fasting plasma glucose level of $\geq 5.0$ mmol/L, 1h level of $\geq 10.0$ mmol/L or 2h level of $\geq 8.5$ mmol/L (IADPSG criteria) | GDM diagnosed following universal screening with a two-step strategy whereby women with a positive glucose challenge screen result (1h venous plasma glucose level $\geq 8.0$ mmol/L) proceeded to a 2h 75g OGTT. GDM diagnosed in presence of either a fasting plasma glucose level of $\geq 5.5$ mmol/L or 2h level of $\geq 8.0$ mmol/L |
| **Predictors** | Age, BMI, family history of diabetes, past history of GDM, history of poor obstetric outcome<sup>b</sup> and ethnicity categories<sup>c</sup> | Age, BMI, ethnicity (8 categories based on self-identified ethnicity), family history of diabetes, past history of GDM and history of poor obstetric outcome |
| **Missing data** | Missing data imputed using multiple imputation by chained equations | Complete case analysis (ie cases with missing data were excluded) |

Abbreviations: GDM, gestational diabetes mellitus; OGTT, oral glucose tolerance test; IADPSG, International Association of Diabetes and Pregnancy Study Groups; BMI, body mass index.
<sup>a</sup> The Original Monash Model for the prediction, in early pregnancy, of the diagnosis of gestational diabetes mellitus was developed and reported in 2011.<sup>7</sup>
<sup>b</sup> History of poor obstetric outcome present if positive for any one of past history of delivery of macrosomic baby, past history of shoulder dystocia or past history of preeclampsia or eclampsia.
<sup>c</sup> Models A, B and C1 with 8 ethnicity categories; Model C2 with 6 ethnicity categories.

**Table 2:** Distribution of patient characteristics in the original development dataset and the 2021 validation and updating dataset (original and imputed sample) and the proportion with missing data.

|  | Original 2011 development dataset <sup>a</sup> (n = 2880) | 2021 Validation and updating dataset |  |
| --- | --- | --- | --- |
|  |  | Original sample (n = 26474) | Imputed sample <sup>a</sup> (n = 26474) |
| Variable | n (%) | n (%) | n (%) |
| Social / demographic factors |  |  |  |
| Age (years) |  |  |  |
| <25 | 396 (13.8) | 3626 (13.7) | 3626 (13.7) |
| 25-29 | 853 (29.6) | 7665 (28.95) | 7665 (28.95) |
| 30-34 | 908 (31.5) | 9559 (36.11) | 9559 (36.11) |
| 35-39 | 568 (19.7) | 4532 (17.12) | 4532 (17.12) |
| ≥40 | 155 (5.4) | 1092 (4.12) | 1092 (4.12) |
| Ethnicity |  |  |  |
| Ethnicity eight categories |  |  |  |
| Anglo-Australian | 1234 (42.8) | 2613 (9.87) | 8262 (31.21) |
| Polynesian | 50 (1.7) | 341 (1.29) | 939 (3.55) |
| Mainland SE Asian | 295 (10.2) | 376 (1.42) | 871 (3.29) |
| Maritime SE Asian | 117 (4.1) | 100 (0.38) | 291 (1.1) |
| Chinese Asian | 189 (6.6) | 616 (2.33) | 1474 (5.57) |
| Southern Asian | 341 (11.8) | 2939 (11.1) | 5505 (20.79) |
| African | 135 (4.7) | 552 (2.09) | 1964 (7.42) |
| Other | 512 (17.8) | 2496 (9.43) | 7168 (27.08) |
| Missing | 0 (0) | 16441 (62.1) | 0 <sup>d</sup> |
| Ethnicity six categories <sup>b</sup> |  |  |  |
| Caucasian | NA | 12098 (45.7) | 12098 (45.7) |
| Southern and Central Asian | NA | 3540 (13.37) | 3540 (13.37) |
| South-East and North-East Asian | NA | 1685 (6.36) | 1685 (6.36) |
| North African, Middle Eastern & Sub-Saharan African | NA | 2240 (8.46) | 2240 (8.46) |
| Oceanian not Australian | NA | 154 (0.58) | 154 (0.58) |
| Other | NA | 6757 (25.52) | 6757 (25.52) |
| missing | NA | 0 <sup>d</sup> | 0 <sup>d</sup> |
| Obstetric and family history |  |  |  |
| Family history of diabetes |  |  |  |
| No | 1735 (60.2) | 14416 (54.45) | 15281 (57.72) |
| Yes | 1145 (39.8) | 10518 (39.73) | 11193 (42.28) |
| missing | 0 (0) | 1540 (5.82) | 0 <sup>d</sup> |
| Past history of GDM <sup>c</sup> |  |  |  |
| No | 2826 (98.1) | 19974 (75.45) | 24611 (92.96) |
| Yes | 54 (1.9) | 1705 (6.44) | 1863 (7.04) |
| missing | 0 (0) | 4795 (18.11) | 0 <sup>d</sup> |
| History of poor obstetric outcome <sup>e</sup> |  |  |  |
| No | 2777 (96.4) | 23384 (88.33) | 25245 (95.36) |
| Yes | 103 (3.6) | 1143 (4.32) | 1229 (4.64) |
| missing | 0 (0) | 1947 (7.35) | 0 <sup>d</sup> |
| Physical characteristics |  |  |  |
| Pre-pregnancy body mass index, kg /m<sup>2</sup> |  |  |  |
| <20.0 | 331 (11.4) | 2434 (9.19) | 2447 (9.24) |
| 20.0–24.9 | 1129 (39.2) | 10669 (40.3) | 10704 (40.43) |
| 25.0–26.9 | 279 (9.7) | 3563 (13.46) | 3572 (13.49) |
| 27.0–29.9 | 266 (9.2) | 3915 (14.79) | 3933 (14.86) |
| 30.0–34.9 | 193 (6.7) | 3353 (12.67) | 3365 (12.71) |
| ≥35.0 | 231 (8.0) | 2442 (9.22) | 2453 (9.27) |
| missing | 451 (15.7) | 98 (0.37) | 0<sup>d</sup> |
| **Diagnosis of gestational diabetes mellitus<sup>f</sup>** | 250 (8.9%) | 4756 (17.96) | 4756 (17.96) |

<sup>a</sup> The Original Monash Model for the prediction, in early pregnancy, of the diagnosis of gestational diabetes mellitus was developed and reported in 2011.<sup>7</sup>
<sup>b</sup> Where missing, deduced using proxy information; country of birth and preferred language.
<sup>c</sup> Set at zero if parity was zero.
<sup>d</sup> Imputed using Multiple Imputation by Chained Equations (MICE) algorithm.
<sup>e</sup> History of poor obstetric outcome present if positive for any one of past history of delivery of macrosomic baby, past history of shoulder dystocia or past history of preeclampsia or eclampsia.
<sup>f</sup> Gestational diabetes mellitus was diagnosed in the Original 2011 model development dataset with a two-step screening strategy and modified WHO criteria and in the 2021 validation and updating dataset with a one-step universal screening strategy and International Association of Diabetes and Pregnancy Study Groups criteria.
Abbreviations: NA, not available as not reported in the publication that first reported the development of the Monash Model.<sup>7</sup>

Updated six-category ethnicity system reduced the number of women assigned to the ‘other’ category as compared to the original eight-category system. Missing data is in Table 2 and was most notable for ethnicity. Ethnicity was deduced for 16,774 women using country of birth and preferred language, both of which had no missing data. In this imputed sample 44.9% were Caucasian, 28.9% Southern and Central Asian, 13.0% South East or North East Asian, and 10.0% North African, Middle Eastern or Sub-Saharan African.

#### Step 1: Performance of the Original Model when temporally validated

Performance of the Original Model to predict GDM in the new dataset is reported in Table 3. Discrimination was reasonable (*c*-statistic of 0.698, 95% CI: 0.690-0.707). However, the calibration plot demonstrated the predicted risks are too extreme

**Table 3:** Coefficients and performance of the Original Model for diagnosis of gestational diabetes when externally validated in validation and updating dataset and Updated Models A, B and C.

| Coefficients | Original model <sup>a</sup> when externally validated | Updated Model A | Updated Model B | Updated Model C1 | Updated Model C2 |
| --- | --- | --- | --- | --- | --- |
| Age (years) |  |  |  |  |  |
| <25 | 1 | 1 | 1 | 1 | 1 |
| 25-29 | 0.92 | 0.92 | 0.57 | 0.33 | 0.29 |
| 30-34 | 1.22 | 1.22 | 0.76 | 0.60 | 0.54 |
| 35-39 | 1.69 | 1.69 | 1.05 | 0.81 | 0.77 |
| ≥40 | 1.95 | 1.95 | 1.21 | 1.01 | 1.01 |
| Body mass index (kg /m<sup>2</sup>) |  |  |  |  |  |
| <20.0 | 1 | 1 | 1 | 1 | 1 |
| 20.0–24.9 | 0.53 | 0.53 | 0.33 | 0.28 | 0.27 |
| 25.0–26.9 | 0.69 | 0.69 | 0.43 | 0.59 | 0.59 |
| 27.0–29.9 | 0.83 | 0.83 | 0.52 | 0.87 | 0.87 |
| 30.0-34.9 | 1.28 | 1.28 | 0.80 | 1.00 | 1.05 |
| ≥35.0 | 1.82 | 1.82 | 1.13 | 1.49 | 1.64 |
| Ethnicity, eight categories |  |  |  |  |  |
| Anglo-Australian | 1 | 1 | 1 | 1 | n/a |
| Polynesian | 1.03 | 1.03 | 0.64 | 0.39 | n/a |
| Mainland SE Asian | 1.61 | 1.61 | 1.00 | 1.12 | n/a |
| Maritime SE Asian | 1.13 | 1.13 | 0.70 | 0.75 | n/a |
| Chinese Asian | 1.31 | 1.31 | 0.82 | 1.07 | n/a |
| Southern Asian | 1.03 | 1.03 | 0.64 | 0.80 | n/a |
| African | 0.69 | 0.69 | 0.43 | 0.82 | n/a |
| Other | 0.18 | 0.18 | 0.11 | 0.79 | n/a |
| Ethnicity, six categories |  |  |  |  |  |
| Caucasian | NA | NA | NA | N/A | 1 |
| Southern and Central Asian | NA | NA | NA | N/A | 1.07 |
| SE and NE Asian | NA | NA | NA | N/A | 1.26 |
| North African, Middle Eastern & Sub-Saharan African | NA | NA | NA | N/A | 0.76 |
| Oceanian not Australian | NA | NA | NA | N/A | 0.43 |
| Other | NA | NA | NA | N/A | 0.90 |
| Family history of diabetes |  |  |  |  |  |
| No | 1 | 1 | 1 | 1 | 1 |
| Yes | 0.53 | 0.53 | 0.33 | 0.39 | 0.38 |
| Past history of GDM |  |  |  |  |  |
| No | 1 | 1 | 1 | 1 | 1 |
| Yes | 2.39 | 2.39 | 1.49 | 1.49 | 1.48 |
| Poor obstetric outcome |  |  |  |  |  |
| No | 1 | 1 | 1 | 1 | 1 |
| Yes | 0.26 | 0.26 | 0.16 | 0.13 | 0.14 |
| Intercept | -5.31 | -4.75 | -3.38 | -3.63 | -3.58 |
| **Model Performance** |  |  |  |  |  |
| C-statistic | 0.6984<br>(0.6901-0.7068) | 0.6984<br>(0.6901-0.7068) | 0.6984<br>(0.6901-0.7068) | 0.7190<br>(0.7110-0.7270) | 0.7324<br>(0.7246-0.7402) |
| Calibration-in-the-large | 0.0858 | 0.0674 | 0.0010 | 0.0027 | 0.0014 |
| Calibration slope | 0.8414 | 0.6890 | 0.9943 | 0.9848 | 0.9920 |
| E:O | 0.4846<br>(0.4819-0.4873) | 0.7550<br>(0.7519-0.7581) | 1.0048<br>(1.0039-1.0056) | 1.0289<br>(1.0279-1.0300) | 1.0212<br>(1.0202-1.0222) |
| Log-likelihood | -12357 | -11713 | -11345 | -11171 | -11038 |
| AIC | 24758 | 23470 | 22734 | 22386 | 22116 |

<sup>a</sup> The Original Monash Model for the prediction, in early pregnancy, of the diagnosis of gestational diabetes mellitus was developed and reported in 2011.<sup>7</sup>
Abbreviations: SE, South East; NE, North East; GDM, gestational diabetes mellitus; CI, confidence interval. E:O, Expected to observed ratio.
AIC = $-2(\log\text{-likelihood}) + 2k$ , where k is number of parameters in model.

#### Step 2: Updated Models and comparisons

Performance of the Updated Models are compared to each other and the Original Model in Table 4. Calibration plots are in Figure 2.

**Table 4:** Model fit comparison.

**(A) Comparison of Models by examining AIC values**
| *The AIC rule of thumb, outlined e.g. in Burnham & Anderson 2004 (if AIC difference is >10 then there is no support to select the model with the larger AIC and the model with the lower AIC is the preferred model with better fit). |  |  |
| --- | --- | --- |
|  | AIC difference | Preferred model * |
| C1 vs C2 | -270 | C2 |
| Original model vs C2 | -2642 | C2 |
| Model A vs C2 | -1354 | C2 |
| Model B vs C2 | -618 | C2 |

**(B) Comparison of Models by examining Log-likelihood scores**
|  | Difference in Log-likelihood | p-value<sup>a</sup> |
| --- | --- | --- |
| C1 vs C2 | 266 | <0.0001 |
| Original model vs C1 | 2372 | <0.0001 |
| Model A vs C1 | 1083 | <0.0001 |
| Model B vs C1 | 347 | <0.0001 |
| Original model vs C2 | 2638 | <0.0001 |
| Model A vs C2 | 1349 | <0.0001 |
| Model B vs C2 | 614 | <0.0001 |

**Figure 2.**
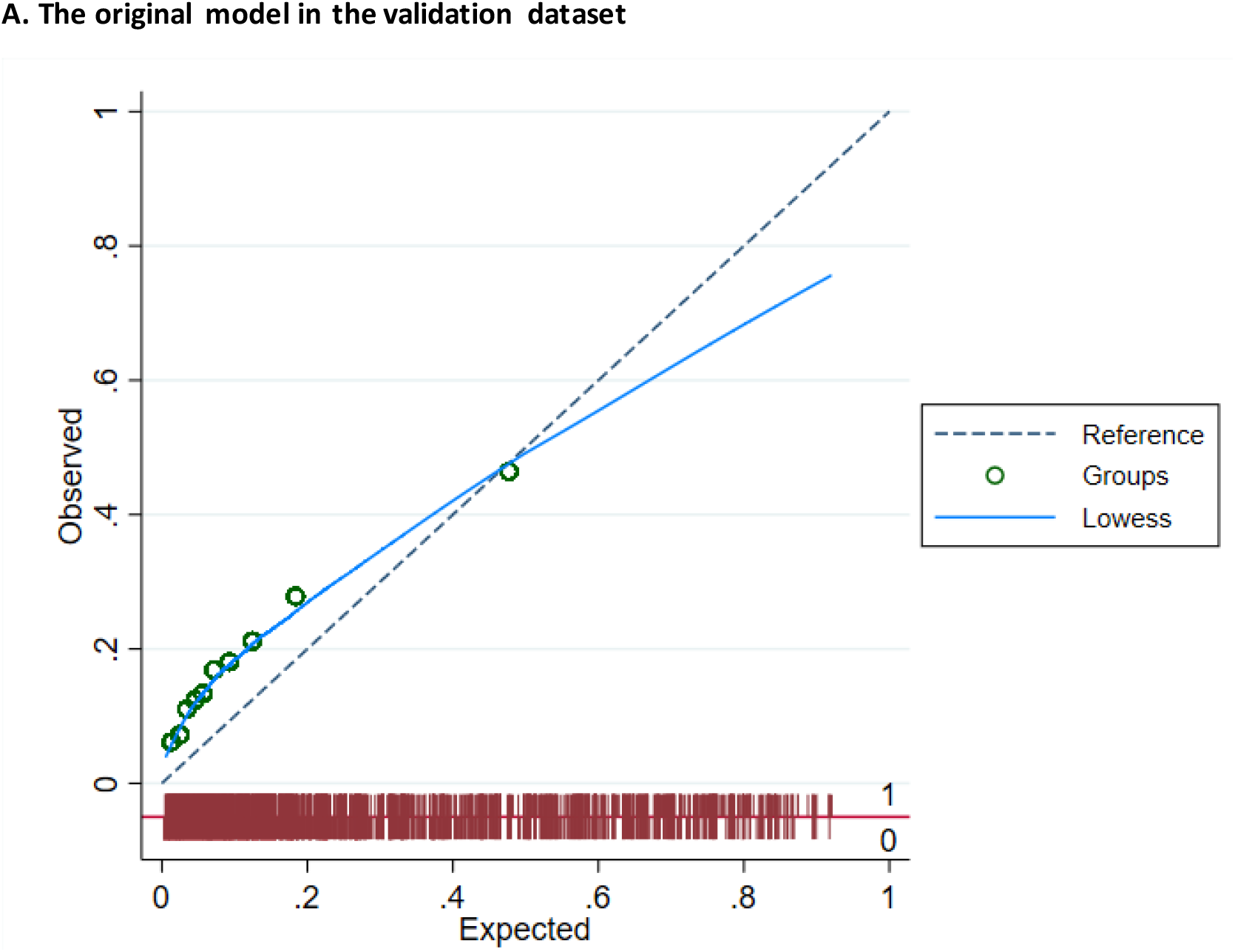
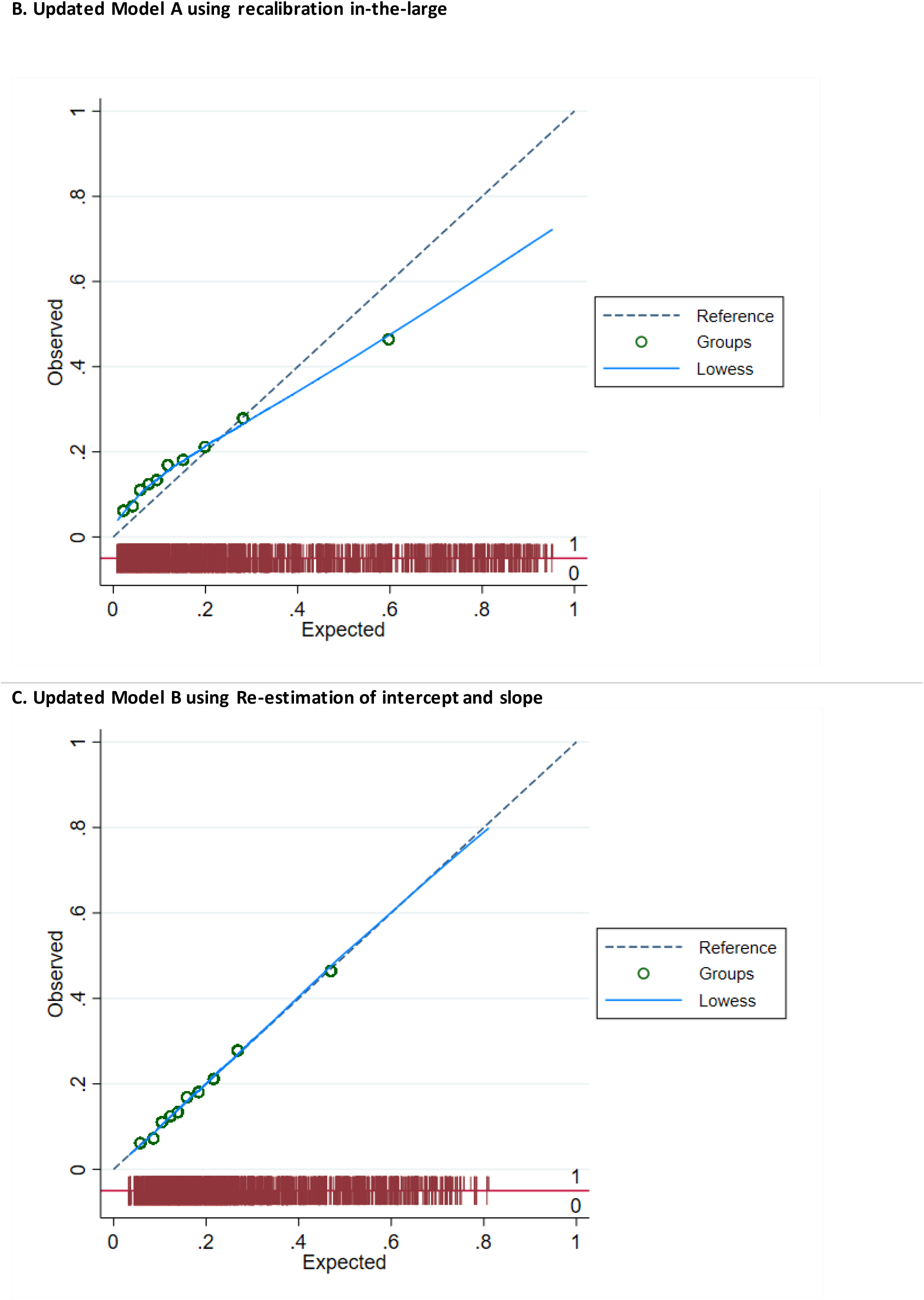
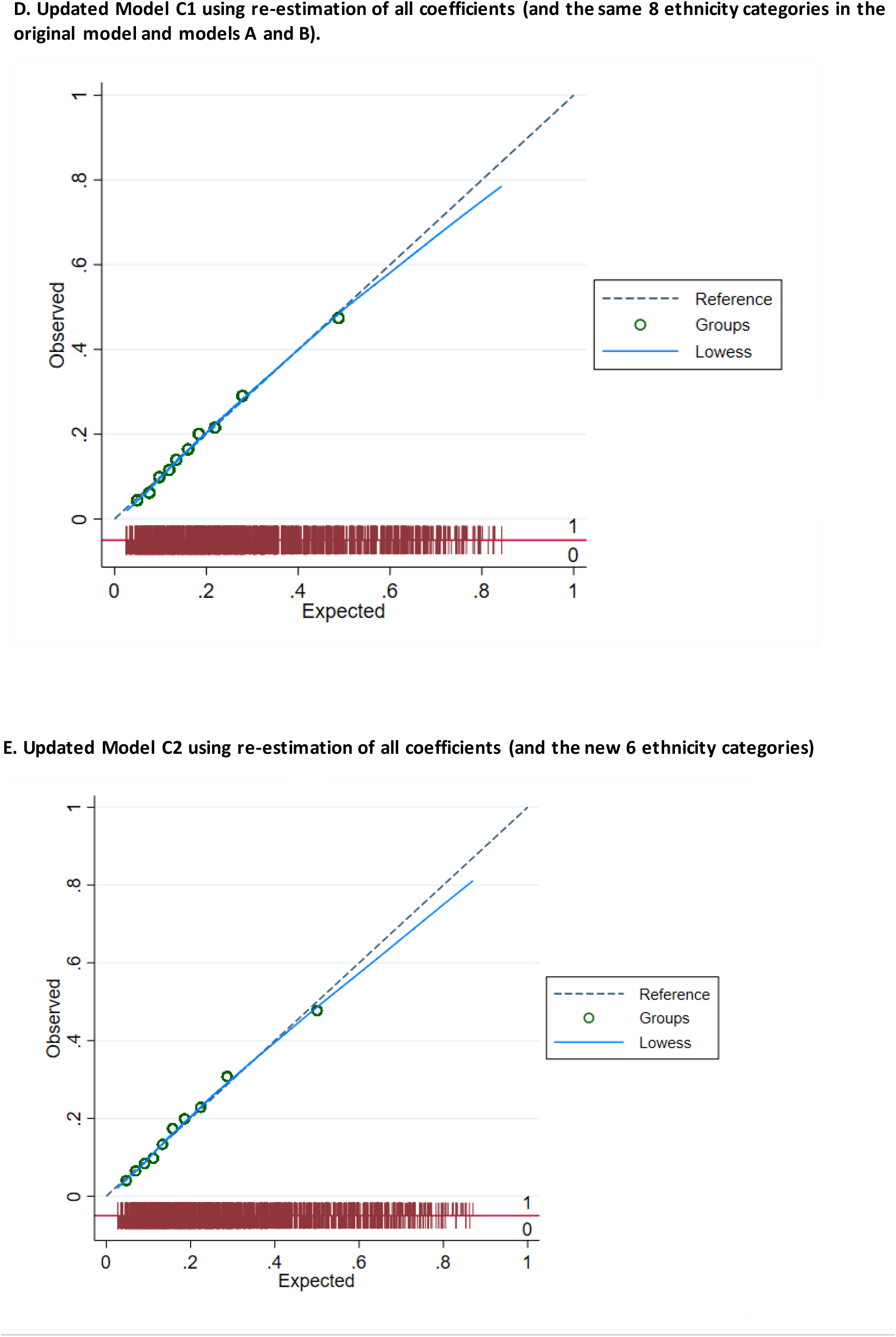
**Calibration plots comparing observed versus predicted risk of diagnosis of gestational diabetes of the original model in the validation dataset (a), and Updated Models (b-e). The plots are grouped by tenths of the predicted risk (circle) and supplemented by a smoothed (Lowess) line. A spike plot of the distribution of events (diagnosis of gestational diabetes mellitus) and non-events (red). Perfect predictions should lie on the 45 º reference (dashed).**

The c-statistic was 0.698 (95% CI: 0.690-0.707) for three models: Original Model and Models A and B. Discrimination improved for Model C1 to 0.719 (95% CI: 0.711-0.727) and further for Model C2 to 0.732 (95% CI: 0.725-0.740), see Table 3.

The calibration plot demonstrating the best aligned calibration plot (closely following 45 degree line) was Model B (Figure 2), showing excellent agreement between predicted and observed risks with a calibration slope of 0.994 and calibration-in-the-large of 0.001 (Table 3). Models C1 and C2 also showed excellent calibration, as reflected in calibration slopes of 0.985 and 0.992 respectively and calibration-in-the-large of 0.003 and 0.001, respectively. Models C1 and C2 calibration plots showed excellent agreement over the majority of the predicted probability range, with the exception of slight over estimation of risk at the upper end (Figure 2). However, most women produced probabilities for GDM using Models C1 and C2 in the well calibrated region (mid-to-low risk) as 96% of women had a risk probability of <0.5.

E:O ratios showed poor agreement for the Original Model and Model A with E:O ratios of 0.48 (95% CI: 0.48-0.49) and 0.76 (0.75-0.76) respectively (Table 3). Excellent E:O ratios were evident with Models B, C1 and C2 at 1.00 (1.00-1.01); 1.03 (1.03-1.03); and 1.02 (1.02-1.02), respectively.

Inspection of the log-likelihood scores and AIC values for each model (Tables 3 and 4) indicated that Models B, C1, and C2 fitted the data better than the Original Model and Model A, as lower scores are associated with better fit. The closed-loop testing procedure comparing log-likelihood scores showed Model C2 having the better fit, which was significantly different to all other models (original, A, B, C1).

## Discussion

Here we temporally validate performance and update the Original Model of the Monash early pregnancy prediction for GDM diagnosis. We use large validation and updating datasets derived from a contemporary, ethnically diverse population with GDM diagnosis based on IADPSG criteria and a universal screening strategy, with a GDM prevalence of 18.0%. We applied best practice in ongoing prediction tool updating. First, we evaluated and temporally validated the Original Model which demonstrated acceptable discrimination, but calibration was not optimal, limiting the accuracy of predicted risks. Next, we updated the model using three different methods with progressively greater variation from the Original Model. Model C2 included a six-category self-identified ethnicity system and displayed very good discrimination, while benefitting from a substantial improvement in calibration. Finally, we compared the updated models to select the best option which best balances improved performance with the risk of overfitting to the validation and updating datasets, and losing the value of the Original Model, which has been independently externally validated. Our results show good model performance for Models B, C1 and C2 when examining discrimination and calibration measures (c-statistic, Calibration-in-the-large, Calibration slope, E:O), demonstrating that with a sufficiently large validation sample size (>25,000 women) the Model B approach to recalibrate the coefficients is very good and converges to similar coefficients generated by the Models C1 and C2 approach. Model B also appeared to show the overall better calibration plot, whilst Model C2 had the better c-statistic. These findings show that our final models (Models B, C1 and C2) are robust, and not likely to be limited by overfitting. Overall, Model C2 is selected as our preferred model because of the comparable calibration plot in the high prevalence region (mid to low risk is associated with the majority of women, i.e. 96% of women in our sample had a risk probability of 0.5 or less), the superior c-statistic (0.73), the more generalizable 6 ethnicity categories and displayed significantly better fit in closed testing (log-likelihood scores and AIC values).

The ultimate goal of developing a clinical prediction model is to integrate it into clinical practice such that an individual’s absolute probability of a future outcome can be used to guide and improve clinical care. There are multiple potential use cases dependent upon the specific clinical needs of the local setting. Firstly, the model can be used in settings where selective GDM screening is routine, to more precisely and accurately identify women for OGTT screening who are at higher risk of developing GDM, than the prevailing single risk-factor based approaches. Secondly, this model can be applied in settings where universal screening is routine, such as Australia, to adapt to disaster settings such as limiting OGTT’s during the COVID-19 pandemic.^11^ Thirdly, this diagnostic model can identify high risk women for targeted preventative lifestyle interventions in past and current clinical trials ^29^ such as the Bump2Baby and Me trial^30^ or ultimately as part of routine care. Finally, it may be used to create a risk-stratified antenatal care pathway, which may be associated with improved perinatal outcomes and cost reduction.^31^

Given the controversy and lack of consistency in GDM screening strategies and diagnostic criteria, clinical prediction models for GDM need to be broadly validated in populations and contexts with variability in these factors. Methodologically, validation studies are important prior to implementation. The current study demonstrates that the Original Model had a reasonable c-statistic but the calibration was problematic and overall did not perform well in the new population diagnosed using IADPSG criteria. However, three updated models (B, C1 and C2) performed better, in the population diagnosed using the IADPSG criteria. The current IADPSG criteria are endorsed by the WHO,^32^ adopted in many regions including within a universal screening strategy in Australia,^23^ and within a selective screening strategy as is common in Europe.^33^ The recent external validation of the Original Model done in the Netherlands provides confidence that the predicator variables used in this study performs well in populations diagnosed with GDM using IADPSG criteria, whether by selective or universal screening.^10^

The work confirms that model updating, retaining original selected variables, via complete model revision where all coefficients are re-estimated using a large (>25,000) diverse multicultural population dataset, improves both discrimination and calibration of the Original Model. The work also demonstrates that model updating using a large and diverse real-world routine health dataset can produce results similar to the best possible model using the Method B approach ^17^. In our case, similar outputs of Models B, C1 and C2 provide evidence that overfitting it unlikely to have occurred in our best model (Model C2), which is extremely important to demonstrate before any clinical utility of the model can be explored (i.e. using this stepped approach in this way is novel and provides important insights into development of robust risk prediction models in large diverse data sets). Noted however is that the methods to undertake Model A and B approaches involve more complex processes as applied statistical software doesn’t contain a ready to use program and even when using other investigator provided codes, adapting and verification requires a good understanding of expert coding and statistical theory. Therefore in the supplementary file is the codes that we used, and the codes used in Vergouwe, et al. 2017 are provided.

However, when a large diverse data set is unavailable to recalibrate a model to different populations, the approach used to create Models A and B is preferred as it will capitalise on the extensive prior external validation of the Original Model across international settings. This has already been demonstrated in examples with data sets less than n=3000 in Vergouwe, et al. 2017. Our study adds value and advances the field of risk prediction model development by taking this multi-step approach in a large diverse data set, to show that the more parsimonious updating methods such as recalibration-in-the-large or logistic recalibration can lead to improvements in model performance and confidence of not overfitting. Use of a more parsimonious updating method could facilitate presentation of the model with the explicit option to use a setting-specific intercept.^34^ The intercept represents the differences between settings that is not captured by the predictors, providing users the ability to customise the model to their setting, significantly enhancing generalisability. This work is novel, applying recommendations in methodologic clinical prediction modelling literature to a recognised clinical prediction model with established clinical utility. It also has the potential to advance the field by serving as an exemplar for model updating in the future. This is needed especially when population samples consist of non-diverse participants and sample sizes are not large.

The Original Model and the Updated Models can be applied using clinical characteristics that are routinely available in clinical practice thus avoiding the barriers and costs in acquisition of additional information, including biomarkers, an approach that has yielded little additional benefit to date. Model C2 enabled us the opportunity to reclassify ethnicity. Ethnicity is a well-established risk factor for the development of GDM.^15^ However, ethnicity is challenging to classify in contemporary multicultural settings ^35^ and classification systems used in clinical research are often based on country of birth and tend to vary internationally. Here we attempted to create a more accurate and generalisable six-category system and use self-identified ethnicity rather than the surrogate country of birth.

The strengths of this study include the validation of the Original Model using a large population-based dataset from a contemporary ethnically diverse universal healthcare setting. Secondly, the application of methodological advances has advanced the field and shows how updating models can be done. These methods include the use of multiple imputation and validation using ten-fold cross-validation. Finally, we have updated the GDM model with a view to presenting it in a format that is well-suited to contemporary clinical practice. Previously, clinical prediction models were typically presented as simplified risk scores, with the attendant loss of performance, to allow the mental computation of risk estimates by clinicians in the healthcare setting. The ubiquitous availability of mobile devices supports online clinical risk calculators to be readily accessed at the point of care, and has allowed the final model to be presented without simplification here. This is currently in use for the Horizons 2020 EU-funded Bump2Baby and Me international clinical trial on lifestyle intervention in women at risk of GDM.^30^. It is also being adapted into an online clinical risk calculator to identify women at risk of GDM. The calculator is available at https://lifestyle.personalgdm.com.

Limitations included that this Updated Model handles the continuous variables, BMI and age, as categorical variables. It is accepted that this approach can reduce predictive power of a model and may be superceded by electronic risk calculators. However, re-estimating the relationship between BMI and age as continuous variables and the diagnosis of GDM would arguably result in a completely new prediction model and go beyond a validation and updating process. This highlights the pervading tension between externally validating and updating existing models and developing completely new models which in turn then require further external validation to demonstrate generalisability.^36^

In conclusion, this rigorous external validation and model updating study, demonstrates the robustness of this model in early pregnancy, to predict the risk of a diagnosis of GDM across many settings including a population defined by IADPSG criteria within a universal screening strategy and using revised ethnicity categories. We build on the clinical prediction modelling literature to demonstrate the potential value of capitalising on existing validated models by undertaking model updating to sustain predictive performance rather than the *de novo* development of a new model. Clinically, this builds on the accumulating evidence supporting the integration of this model into routine practice to deliver risk-stratified care to women at risk of GDM across a number of clinical use case scenarios and settings.

## Data Availability

All data produced in the present study are available upon reasonable request to the authors

## References

1. Classification and diagnosis of diabetes mellitus and other categories of glucose intolerance. National Diabetes Data Group. Diabetes 1979; 28(12): 1039–57.

2. International Weight Management in Pregnancy Collaborative Group. Effect of diet and physical activity based interventions in pregnancy on gestational weight gain and pregnancy outcomes: meta-analysis of individual participant data from randomised trials. BMJ 2017; 358: j3119.

3. Landon MB, Spong CY, Thom E, et al. A multicenter, randomized trial of treatment for mild gestational diabetes. N Engl J Med 2009; 361(14): 1339–48.

4. Crowther CA, Hiller JE, Moss JR, et al. Effect of treatment of gestational diabetes mellitus on pregnancy outcomes. N Engl J Med 2005; 352(24): 2477–86.

5. Cooray SD, Thangaratinam S, Teede HJ. Prediction modelling to personalise care for gestational diabetes. BJOG 2020.

6. Lamain – de Ruiter M, Kwee A, Naaktgeboren CA, Franx A, Moons KGM, Koster MPH. Prediction models for the risk of gestational diabetes: a systematic review. Diagnostic and Prognostic Research 2017; 1(1): 3.

7. Teede HJ, Harrison CL, Teh WT, Paul E, Allan CA. Gestational diabetes: Development of an early risk prediction tool to facilitate opportunities for prevention. Australian and New Zealand Journal of Obstetrics and Gynaecology 2011; 51(6): 499–504.

8. Thériault S, Forest J-C, Massé J, Giguère Y. Validation of early risk-prediction models for gestational diabetes based on clinical characteristics. Diabetes Research and Clinical Practice 2013; 103(3): 419–25.

9. Lamain-de Ruiter M, Kwee A, Naaktgeboren CA, et al. External validation of prognostic models to predict risk of gestational diabetes mellitus in one Dutch cohort: prospective multicentre cohort study. BMJ 2016; 354: i4338.

10. van Hoorn F, Koster M, Naaktgeboren CA, et al. Prognostic models versus single risk factor approach in first-trimester selective screening for gestational diabetes mellitus: a prospective population-based multicentre cohort study. BJOG 2020.

11. Thangaratinam S, Cooray SD, Sukumar N, et al. ENDOCRINOLOGY IN THE TIME OF COVID-19: Diagnosis and management of gestational diabetes mellitus. Eur J Endocrinol 2020; 183(2): G49–G56.

12. International Association of Diabetes Pregnancy Study Groups Consensus Panel, Metzger BE, Gabbe SG, et al. International association of diabetes and pregnancy study groups recommendations on the diagnosis and classification of hyperglycemia in pregnancy. Diabetes Care 2010; 33(3): 676–82.

13. Wong VW, Lin A, Russell H. Adopting the new World Health Organization diagnostic criteria for gestational diabetes: How the prevalence changes in a high-risk region in Australia. Diabetes Res Clin Pract 2017; 129: 148–53.

14. Cade TJ, Polyakov A, Brennecke SP. Implications of the introduction of new criteria for the diagnosis of gestational diabetes: a health outcome and cost of care analysis. Bmj Open 2019; 9(1): e023293.

15. Yue DK, Molyneaux LM, Ross GP, Constantino MI, Child AG, Turtle JR. Why does ethnicity affect prevalence of gestational diabetesã The underwater volcano theory. Diabet Med 1996; 13(8): 748–52.

16. Collins GS, Reitsma JB, Altman DG, Moons KG. Transparent Reporting of a multivariable prediction model for Individual Prognosis or Diagnosis (TRIPOD): the TRIPOD statement. Ann Intern Med 2015; 162(1): 55–63.

17. Vergouwe Y, Nieboer D, Oostenbrink R, et al. A closed testing procedure to select an appropriate method for updating prediction models. Statistics in Medicine 2017; 36(28): 4529–39.

18. Australian and New Zealand Clinical Trials Registry [Internet]. The Prediction modelling for Risk-Stratified care for women with Gestational Diabetes (PeRSonal GDM) study: Calculating the individualised risk of adverse outcomes for women with gestational diabetes (ACTRN12620000915954). 17 Sep 2020. https://www.anzctr.org.au/ACTRN12620000915954.aspx (accessed 25 Sep 2020).

19. Hoffman L, Nolan C, Wilson JD, Oats JJ, Simmons D. Gestational diabetes mellitus -- management guidelines. The Australasian Diabetes in Pregnancy Society. Med J Aust 1998; 169(2): 93–7.

20. Cooray SD, Boyle JA, Soldatos G, et al. Protocol for development and validation of a clinical prediction model for adverse pregnancy outcomes in women with gestational diabetes. Bmj Open 2020; 10(11): e038845.

21. Janssen KJ, Kalkman CJ, Grobbee DE, Bonsel GJ, Moons KG, Vergouwe Y. The risk of severe postoperative pain: modification and validation of a clinical prediction rule. Anesth Analg 2008; 107(4): 1330–9.

22. Harrell FE, Jr., Lee KL, Mark DB. Multivariable prognostic models: issues in developing models, evaluating assumptions and adequacy, and measuring and reducing errors. Stat Med 1996; 15(4): 361–87.

23. Nankervis A, McIntyre HD, Moses RG, et al. ADIPS Consensus Guidelines for the Testing and Diagnosis of Hyperglycaemia in Pregnancy in Australia and New Zealand. 2014.

24. Steyerberg EW. Clinical Prediction Models: A Practical Approach to Development, Validation, and Updating. Second edition. ed. New York ; London: Springer International Publishing; 2019.

25. Steyerberg EW, Borsboom Gjjm, Van Houwelingen HC, Eijkemans MJC, Habbema JDF. Validation and updating of predictive logistic regression models: a study on sample size and shrinkage. Statistics in Medicine 2004; 23(16): 2567–86.

26. Janssen KJ, Moons KG, Kalkman CJ, Grobbee DE, Vergouwe Y. Updating methods improved the performance of a clinical prediction model in new patients. J Clin Epidemiol 2008; 61(1): 76–86.

27. Australian Bureau of Statistics. 1249.0 - Australian Standard Classification of Cultural and Ethnic Groups (ASCCEG), 2016. 18 July 2016 2016. https://www.abs.gov.au/ausstats/abs@.nsf/mf/1249.0 (accessed 2 October 2019.

28. Steyerberg EW, Vergouwe Y. Towards better clinical prediction models: seven steps for development and an ABCD for validation. Eur Heart J 2014; 35(29): 1925–31.

29. Harrison C, Lombard CB, Strauss BJ & Teede HJ. Optimizing healthy gestational weight gain in women at high risk of gestational diabetes: A randomized controlled trial. Obesity 2013; 21: 904.

30. Bump2Baby and Me. Mother and families | Bump2Baby and Me. 2021. https://bump2babyandme.org/mothers-and-families/ (accessed 18 Mar 2021 2021).

31. van Montfort P, Scheepers HCJ, Dirksen CD, et al. Impact on perinatal health and cost-effectiveness of risk-based care in obstetrics: a before-after study. Am J Obstet Gynecol 2020.

32. Diagnostic criteria and classification of hyperglycaemia first detected in pregnancy: a World Health Organization Guideline. Diabetes Res Clin Pract 2014; 103(3): 341–63.

33. Benhalima K, Mathieu C, Van Assche A, et al. Survey by the European Board and College of Obstetrics and Gynaecology on screening for gestational diabetes in Europe. Eur J Obstet Gynecol Reprod Biol 2016; 201: 197–202.

34. Steyerberg EW. Perioperative Mortality of Elective Abdominal Aortic Aneurysm Surgery. Archives of Internal Medicine 1995; 155(18): 1998.

35. Lockie E, McCarthy E, Hui L, Churilov L, Walker S. Feasibility of using self-reported ethnicity in pregnancy according to the gestation-related optimal weight classification: a cross-sectional study. BJOG: An International Journal of Obstetrics & Gynaecology 2018; 125(6): 704–9.

36. Steyerberg EW, Harrell FE, Jr. Prediction models need appropriate internal, internal-external, and external validation. J Clin Epidemiol 2016; 69: 245–7.

